# Delivering Results: Evaluating the Implementation of Bulamu Healthcare’s Maternal and Newborn Program in Uganda

**DOI:** 10.1101/2025.06.25.25330290

**Authors:** Kylie Dougherty, Andrew Mukuye, Richard Businge, Ziyada Babirye, Samuel David Blizzard, Mackay Masereka, Yvonne Vaucher, Richard Siegler, James Healy, Peter Waiswa, Margaret Nakakeeto, Lisa R. Hirschhorn

**Author notes:** **Corresponding Author**, Kylie Dougherty, Northwestern University Feinberg School of Medicine 625 Michigan Ave., Chicago, IL 60611.

## Abstract

**Background:** Maternal and newborn mortality remain critical challenges in Uganda, with rates exceeding global targets. Despite the existence of effective clinical guidelines, such as WHO’s Essential Newborn Care (ENC) and Helping Mothers Survive (HMS), implementation gaps persist due to training, resource, and systems-level barriers. In response, Bulamu Healthcare launched a comprehensive maternal and newborn care capacity-strengthening program across eight districts in Uganda to improve clinical outcomes and health system performance. This model works in partnership with 104 existing public maternity facilities, which delivered more than 77,000 babies in 2024, and served more than 154,000 mothers and newborns in 2024.

**Methods:** This retrospective evaluation assessed the implementation of Bulamu’s integrated maternal and newborn health model using the RE-AIM framework (Reach, Effectiveness, Adoption, Implementation, and Maintenance). The program included clinical training (WHO ENC, HMS) and mentorship, provision of essential supplies and equipment, and data-driven quality improvement. Data were collected from Uganda’s national District Health Information Software (DHIS2) and Bulamu program monitoring tools from January 2023 to March 2025. Descriptive statistics summarized key implementation and outcome indicators.

**Results:** Bulamu supported 109 health facilities, exceeding the initial target of 79, and established 12 functional neonatal care units. All facilities received the full intervention package. Over 250 maternity workers were trained, and 100% of district health workers in supported areas received ENC training. Live births in Bulamu facilities increased by 14% from 2023 to 2024, while maternal deaths decreased by 59% (from 17 to 7). The proportion of low-birth-weight newborns receiving kangaroo mother care rose from 54% to 62%, and newborn discharged alive rates improved from 80% to 96.4%. Facilities demonstrated early signs of sustainability, with at least one trained maternal and newborn provider retained per site and local staff beginning to independently train peers.

**Conclusions:** Bulamu’s integrated approach combining evidence-based interventions with targeted systems which supports improved care delivery and maternal-newborn outcomes. Findings highlight the utility of the RE-AIM framework for real-world evaluation and support the value of embedded implementation research in strengthening health systems. Future research should examine cost-effectiveness, sustainability, and context-specific implementation mechanisms to inform scale-up and long-term impact.

## 1. Background

Maternal and newborn health is a pressing global challenge, particularly in resource-limited settings. Maternal mortality rates remain persistently high, with a global maternal mortality ratio (MMR) of 224 per 100,000 live births.^1^ In East Africa, the lifetime risk of maternal mortality for women aged 15 and older is approximately 1 in 41, one of the highest in the world.^2^ Furthermore, neonatal mortality is the leading cause of under-five deaths in Africa.^3^ Each year, over three million maternal, neonatal, and stillbirth deaths occur in Africa alone, with more than 75% of these deaths being preventable or treatable through cost-effective interventions.^4,5^ In Uganda, maternal and newborn health indicators reflect these regional challenges. As of 2023, the neonatal mortality rate (NMR) is 22 per 1,000 live births, the stillbirth rate is 18 per 1,000 live births, and the MMR is 189 per 100,000 live births, all of which are far higher than the World Health Organization (WHO) 2030 Sustainable Development Goals (SDG) of NMR less than 12 per 1,000, stillbirth rates less than 10 per 1,000, and MMR less than 70 per 100,000.^6–8^ International organizations, including WHO, the American Academy of Pediatrics, and Jhpiego, have developed effective health worker training programs to address these challenges. Programs such as WHO Essential Newborn Care (ENC),^9^ which includes Helping Babies Breathe^10^, essentials are for every baby, essential care for small babies, and Helping Mothers Survive (HMS)^11^ have demonstrated significant success in reducing maternal and neonatal mortality rates. For example, WHO’s ENC training has been shown to decrease NMR by up to 50%.^9^ Despite their proven effectiveness, these interventions remain underutilized in many low-resource settings, perpetuating preventable deaths.^6^ Barriers to widespread implementation and scaling of these programs—such as limited adoption of WHO ENC, and HMS practices—highlight significant gaps in care.^9,12,13^

Additional factors, such as difficulties for women in accessing healthcare facilities, including delays in transportation or referrals and stockouts of essential supplies, exacerbate the risks faced by mothers and newborns.^14–16^ Studies of Uganda’s health supply chain systems reveal persistent issues with stockouts of critical medicines and supplies, further jeopardizing maternal and newborn care.^17^ Compounding these challenges, delays in receiving adequate treatment after arriving at a healthcare facility—the third delay—remain a critical determinant of adverse maternal and newborn outcomes.^18^ The WHO reported that these challenges are particularly acute in regions like Busoga, Uganda, where delays in reaching healthcare facilities and receiving timely care significantly impact health outcomes.^19^

Recognizing the critical importance of maternal and newborn health, the WHO and other global health stakeholders are calling for interventions that integrate maternal and newborn care, emphasizing the need to begin care during the prenatal period to ensure positive outcomes for both mother and child.^6,20^

In response to these identified challenges preventing high-quality maternal and newborn care, Bulamu Healthcare, a nonprofit organization based in Uganda implemented an integrated maternal and newborn capacity strengthening program in eight districts in Uganda utilizing the above internationally-recommended programs.^21^ Originally, Bulamu was founded in 2016 with aim of strengthening health systems, primary care, and surgery in Uganda. In 2021, Bulamu expanded their work to develop their Health Center Excellence Program that builds upon existing public health facilities and teams to strengthen Uganda’s public health system.^21^ Through the Health Center Excellence Program Bulamu began to support the country’s maternal and child health services at level III and IV facilities and district hospitals. This paper evaluates the implementation success of Bulamu’s maternal and newborn model in Uganda.

## 2. Methods

This study aimed to evaluate Bulamu’s success of implementing their integrated maternal and newborn care capacity-strengthening model across eight districts in Uganda.

### 2.1 Setting and Population

All level III, and IV health centers and district hospitals in eight of the twelve districts throughout the country Bulamu works in were included in this study. The districts supported by Bulamu included: Amuru, Budaka, Bunyangabu, Gulu, Kakumiro, Kyegegwa, Mayuge, and Namutumba. These districts and facilities were selected by Bulamu because of their poor MCH indicator performance at both regional and national level. When evaluating Bulamu-specific outcomes, all pregnant women who sought prenatal care at a Bulamu-supported facility were included in the sample, as well as their newborns.

### 2.2 Bulamu Model

Bulamu’s maternal and newborn care model is a comprehensive approach including evidence-based training and several key strategies.

The trainings included WHO ENC and HMS.^9,11^ To enable the success of this program, Bulamu employed several strategies. First, the training was provided simultaneously to all District unit staff to improve coordination and communication between all District units, after which the training team personally visited all District sites to facilitate implementation. Onsite mentorship was used throughout the clinician training process and intermittently after training was complete. All District clinical services are free of charge, which removes financial barriers to families, and Bulamu provided medical supplies and devices to maternal and newborn care. Furthermore, Bulamu collaborated closely with District and national Ministry of Health leadership to align efforts with national priorities and goals. Data dashboards and maternity client treatment forms were provided to all 109 facilities to support facility and district health teams to perform tracking and quality improvement (QI) initiatives. The belief was that this model of care combined with the implementation strategies would collectively enable high-quality, accessible care for mothers and newborns in Uganda.

### 2.3 Framework

To understand the implementation outcomes, we applied the RE-AIM framework, which encompasses *Reach, Effectiveness, Adoption, Implementation,* and *Maintenance*.^22^ The RE-AIM framework is especially useful when considering holistic evaluation of early-stage programs to ensure they are effectively adopted with fidelity and equity while identifying actionable barriers to implementation. This approach enabled us to assess whether Bulamu’s maternal and newborn care model reached the target population, achieved its intended health outcomes, was widely adopted by healthcare workers, was implemented as planned, and demonstrated potential for long-term sustainability.

To gain a comprehensive understanding of Bulamu’s implementation success, we categorized the RE-AIM outcomes into two key areas: 1) outcomes related to Bulamu’s support activities, and 2) outcomes related to evidence-based interventions (EBIs) and health outcomes. The first category examines how Bulamu provided support to Districts and facilities, including activities like WHO-recommended training, and emergency transportation services. The second category focuses on the impact of Bulamu’s support on health outcomes, such as District coverage rates and percentage changes in practices like kangaroo mother care (KMC). A detailed description of how we operationalized the RE-AIM dimensions for this evaluation can be seen in Table 1.

**Table 1.** Bulamu RE-AIM Outcomes Defined.

| **Bulamu Support Activities** |  |  |
| --- | --- | --- |
| **Dimension** | **Measurement** | **Time Period** |
| **Reach** | Number of healthcare facilities supported by Bulamu | Jan-Dec 2024 |
| **Reach** | Neonatal care units trained and launched | Jan-Dec 2024 |
| **Adoption** | Change in the number of Bulamu-supported sites with a functional neonatal care unit (able to offer services to babies requiring neonatal care) | 2023 vs. 2024 |
| **Implementation-Feasibility** | Number of Bulamu maternity workers trained in WHO essential newborn care | Jan-Dec 2024 |
| **Implementation-Feasibility** | % of district health workers in Bulamu's 8 supported districts who received WHO essential newborn care training | Jan-Dec 2024 |
| **Implementation-Feasibility** | Number of Bulamu expert trainers for WHO essential newborn care trained (ToTs) | Jan-Dec 2024 |
| **Implementation-Feasibility** | Proportion of Bulamu-supported sites that have a functional maternal and neonatal unit (only looks at 2024 coverage) | Jan-Dec 2024 |
| **Implementation-Fidelity** | Proportion of Bulamu-supported sites that received training, medical devices, maternal and newborn supplies, and QI support (looks to see if all facilities received the same intervention) | Jan-Dec 2024 |
| **Maintenance** | Proportion of Bulamu-supported sites with one trained staff member in the maternal unit and one trained staff member in the newborn unit | Jan-Dec 2024 |
| **Maintenance** | Number of Bulamu-supported sites that are independently training other staff | Jan-Dec 2024 |
| **Evidence-based interventions** |  |  |
| **Dimension** | **Measurement** | **Time Period** |
| **Reach** | Number of deliveries conducted in trained facilities (post-training) | Jan-Dec 2024 |
| **Reach** | Percentage change of Bulamu deliveries | 2023 vs. 2024 |
| **Effectiveness** | Percentage change of newborns <2500 at Bulamu sites that were initiated on KMC | 2023 vs. 2024 |
| **Effectiveness** | Percentage of Bulamu postpartum hemorrhage mothers saved | 2024 |
| **Effectiveness** | Percentage change of Bulamu maternal deaths | 2023 vs. 2024 |
| **Effectiveness** | Percentage change of newborns discharged alive for Bulamu sites (number of newborns discharged alive/ total number of small and sick newborns admitted) | 2023 vs. 2024 |
| **Adoption** | Percentage change in referrals from Bulamu sites to other facilities (outside the home district) | 2023 vs. 2024 |
| **Adoption** | Proportion of Bulamu-supported sites that saw an increase in the percentage of newborns <2500 g initiated on KMC | 2023 vs. 2024 |
| **Implementation-Feasibility** | Percentage change in referrals from non-Bulamu supported sites to Bulamu-supported sites | 2023 vs. 2024 |
| **Implementation-Fidelity** | Proportion of newborns <2500 at Bulamu-supported sites that were initiated on KMC | Jan-Dec 2024 |
| **Maintenance** | Proportion of newborns <2500 at Bulamu-supported sites that were initiated on KMC | Jan-March 2025 |
| **Maintenance** | Proportion of women of women at Bulamu-supported sites who received uterotonic in 3<sup>rd</sup> stage of labor | Jan-March 2025 |

### 2.4 Data Collection

All mortality data and newborn care indicators were extracted from Uganda’s District Health Information Software (DHIS2), a nationwide online dashboard of health outcomes by region and facility for the period January 2023 through December 2024.^23^ Program outcomes, such as staff trained, number of live deliveries, number of children discharged alive, and functional status of equipment, were tracked by Bulamu team members and outcomes were extracted from the routine programmatic data. Bulamu-supported staff used the maternity client treatment and neonatal unit admission forms to routinely report health outcomes, including the provision of KMC. These were captured in the Maternity and Neonatal registers respectively. Finally, referrals to and out of Bulamu-supported sites were collected from the referral tracker/Register.

### 2.5 Data Analysis

Data analysis was performed in April 2025 using. The data included [list types of data, e.g., antenatal care attendance, skilled birth attendance, immunization coverage, etc.], disaggregated by [e.g., district, health facility, month]. Data was cleaned and analyzed using the built-in DHIS2 analytics tools, including the Data Visualizer and Pivot Table applications, to generate descriptive statistics and visualize trends over time.

Where necessary, data were exported to Microsoft Excel for further aggregation and analysis. Indicators were calculated based on standard national definitions and WHO guidelines. Data completeness and consistency were assessed prior to analysis to ensure validity and reliability.

Descriptive statistics were used to report implementation outcomes and differences in coverage rates over time.

## 3. Results

This section presents findings from the implementation of Bulamu’s maternal and newborn care model across eight districts in Uganda, organized using RE-AIM. Results are grouped into two categories: 1) outcomes related to Bulamu’s support activities and 2) outcomes related to EBIs and health outcomes. Unless otherwise noted, data were collected from January-December in 2023 and 2024, with select *maintenance* outcomes measured through March 2025. Table 2 presents the complete RE-AIM outcomes.

**Table 2.** Bulamu RE-AIM Outcomes.

| Dimension | Measurement | Time Period | Outcome |
| --- | --- | --- | --- |
| **Reach** | Number of healthcare facilities supported by Bulamu | Jan-Dec 2024 | 8/8 Districts, 109/79 facilities<br>*Target was 79, but identified facilities that had been upgraded from level 2 to level 3* |
| **Reach** | Neonatal care units trained and launched | Jan-Dec 2024 | 12/12 units |
| **Adoption** | Change in number of Bulamu-supported sites with a functional neonatal care unit (able to offer services to babies requiring neonatal care) | 2023 vs. 2024 | 2023-no functional NCU<br>2024-12/13 |
| **Implementation-Feasibility** | Number of maternity workers trained in WHO essential newborn care | Jan-Dec 2024 | 256 maternity workers |
| **Implementation-Feasibility** | % of district health workers in Bulamu's 8 supported Districts who received WHO essential newborn care training | Jan-Dec 2024 | 100% |
| **Implementation-Feasibility** | Number of expert trainers for WHO essential newborn care trained (ToTs) | Jan-Dec 2024 | 36 trainers |
| **Implementation-Feasibility** | Proportion of Bulamu-supported sites that have a functional maternal and neonatal units (only looks at 2024 coverage) | Jan-Dec 2024 | 12/12 (100%) |
| **Implementation-Fidelity** | Proportion of Bulamu-supported sites that received training, medical devices, maternal and newborn supplies, and QI support (looks to see if all facilities received the same intervention) | Jan-Dec 2024 | 100% |
| **Maintenance** | Proportion of Bulamu-supported sites with one trained staff member in the maternal unit and one trained staff member in the newborn unit | Jan-Dec 2024 | 100% |
| **Maintenance** | Proportion/number of Bulamu-supported sites that are independently training other staff (ex. Budaka) | Jan-Dec 2024 | 1/12 (Budaka HC IV) |

**Primary Outcomes: Evidence-Based Intervention (ANC, resuscitation, delivery etc.)** **Implementation RE-AIM** (*This explores what Bulamu's support did for health outcomes*)
| **Dimension** | **Measurement** | **Time Period** | **2023** | **2024** | **2023 – 2024 Change (n, %)** |
| --- | --- | --- | --- | --- | --- |
| **Reach** | Number of deliveries conducted in trained facilities (post-training) | Jan-Dec 2024 | --- | 76523 deliveries | --- |
| **Reach** | Percentage change of Bulamu deliveries | 2023 vs. 2024 | 68,367 | 76,523 | 8,156 (+11.9%) |
| **Effectiveness** | Percentage change of newborns <2500 at Bulamu sites that were initiated on KMC | 2023 vs. 2024 | 2023: 54% | 2024: 62% | Change: +8% |
| **Effectiveness** | Percentage of postpartum hemorrhage mothers saved | 2023 vs 2024 | 2023<br>612 of 620 women (98.7%) | 2024<br>649 of 651 women (99.6%) | Change of +1% |
| **Effectiveness** | Percentage change of maternal deaths | 2023 vs. 2024 | 2023 maternal deaths (17) | 2024 maternal deaths (7) | Change - 59% |
| **Effectiveness** | Percentage change of newborns discharged alive for Bulamu sites | 2023 vs. 2024 | Start -80% (201/252) | End- 96.4% (3,727/3,866) | Change 16.4% |
| **Adoption** | Percentage change in referrals from Bulamu sites to higher level facilities outside the home District) | 2023 vs. 2024 | 1,775 referrals out in 2023 | 1,648 referrals out in 2024 | Change: -7% |
| **Adoption** | Proportion of Bulamu-supported sites that saw an increase in the percentage of newborns <2500 g initiated on KMC | 2023 vs. 2024 | --- | 45/109 (41.3%) | --- |
| **Implementation -Feasibility** | Percentage change in referrals from non-Bulamu supported Districts to Bulamu-supported District | 2023 vs. 2024 | 4228 referrals in 2023 | 4539 referrals in 2024 | Change: +7% |
| **Implementation - Fidelity** | Proportion of newborns <2500 grams at Bulamu-supported sites that were initiated on KMC | Jan-Dec 2024 | --- | 62% | --- |
| **Maintenance** | Proportion of newborns <2500 grams at Bulamu-supported sites that were initiated on KMC | Jan-April 2025 | --- | 68% | --- |
| **Maintenance** | Proportion of women of women who received uterotonic in 3<sup>rd</sup> stage of labor | Jan-April 2025 | --- | 95% | --- |

### 3.1 RE-AIM for Bulamu support activities

Bulamu’s maternal and newborn health program successfully *reached* its target population, by supporting all eight targeted districts and 109 facilities (104 public facilities and 5 private facilities) in eight districts. Although the original implementation goal was to reach 79 facilities, additional health centers were included in the program due to facility upgrades from level II to level III. *Reach* was also demonstrated through full District engagement and development of 12 new neonatal care units which were trained and launched during 2024, representing 100% coverage of planned neonatal care unit support.

*Adoption* of the program was high, with a notable increase in the proportion of Bulamu-supported sites operating functional neonatal care units capable of caring for newborns requiring specialized attention. In 2023, 3 facilities had non-functional units and 9 had no units at all capable of delivering specialized care for newborns. However, by 2024 all targeted sites (12 facilities) had an operational newborn care unit for small and sick newborns. *Implementation feasibility* was further evidenced by the training of 256 maternity workers in ENC. This resulted in the successful training of 100% of district health workers across all eight Bulamu-supported Districts. Additionally, 36 expert trainers were prepared through a training-of-trainers (ToT) model, ensuring local capacity to support future scale-up and training needs. Furthermore, all facilities had functional maternal and neonatal units in place by the end of 2024, demonstrating the full *feasibility* of establishing operational service delivery platforms within the public health system.

Program *implementation fidelity* was strong, with all supported facilities receiving the full intervention package, including staff training, essential medical devices, and maternal and newborn care supplies. This consistency in delivery ensured that all sites benefited equally from the program’s intended components. In terms of *maintenance,* every Bulamu-supported facility retained at least one trained staff member in both maternal and newborn care units as of March 2025. Notably, Budaka Health Center IV began independently training additional staff without direct program involvement, demonstrating early signs of sustainable capacity-building within the system.

### 3.2 RE-AIM for EBIs and health outcomes

The implementation of Bulamu’s support package also contributed to improved clinical service delivery and health outcomes across supported sites. In terms of *reach,* a total of 76,523 deliveries were conducted in Bulamu-supported facilities in 2024, representing a 12% increase from 2023 when 68,367 deliveries were recorded (Figure 2). Several *effectiveness* indicators also improved over the course of the program. The proportion of newborns weighing less than 2500 grams who were initiated on KMC rose from 54% in 2023 to 62% in 2024 (Figure 3). Maternal deaths decreased from 17 in 2023 to 7 in 2024, representing a 59% reduction. The percentage of newborns discharged alive improved from 80% at baseline to 96.4% by the end of 2024.

**Figure 1.**
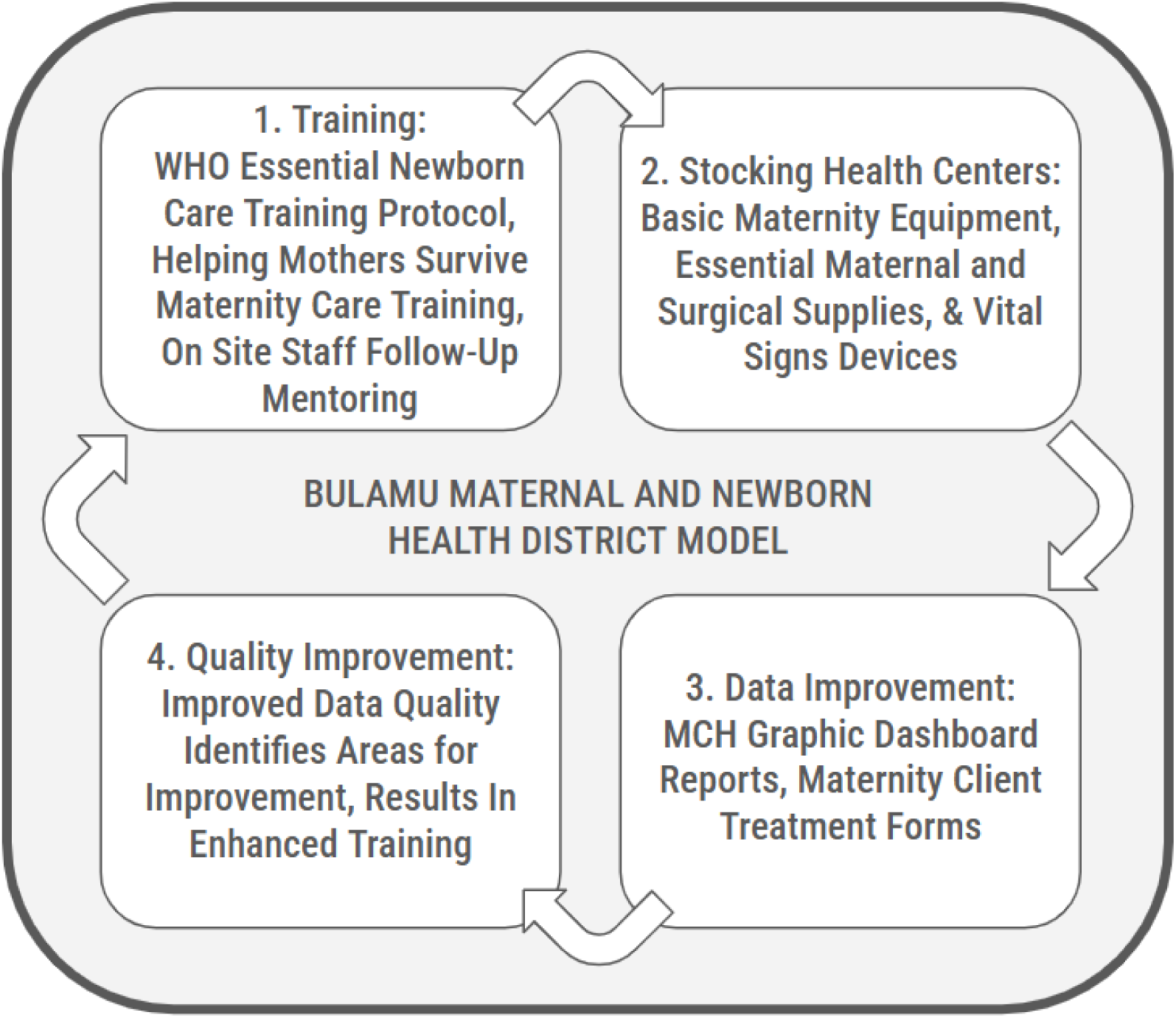
Description of Bulamu maternal and newborn care model

**Figure 2.**
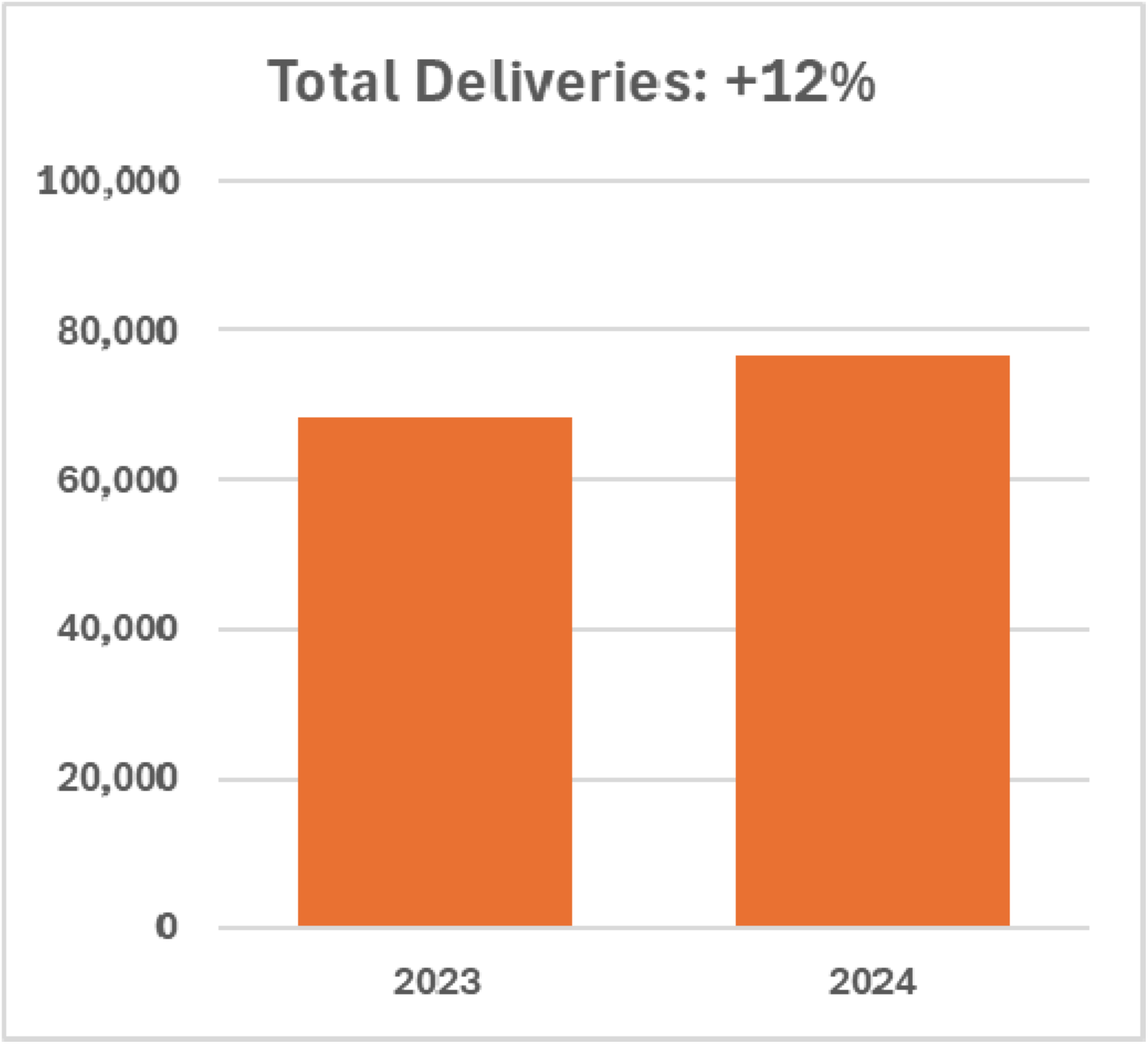
Total number of delivered within Bulamu-supported districts in 2023 and 2024

**Figure 3.**
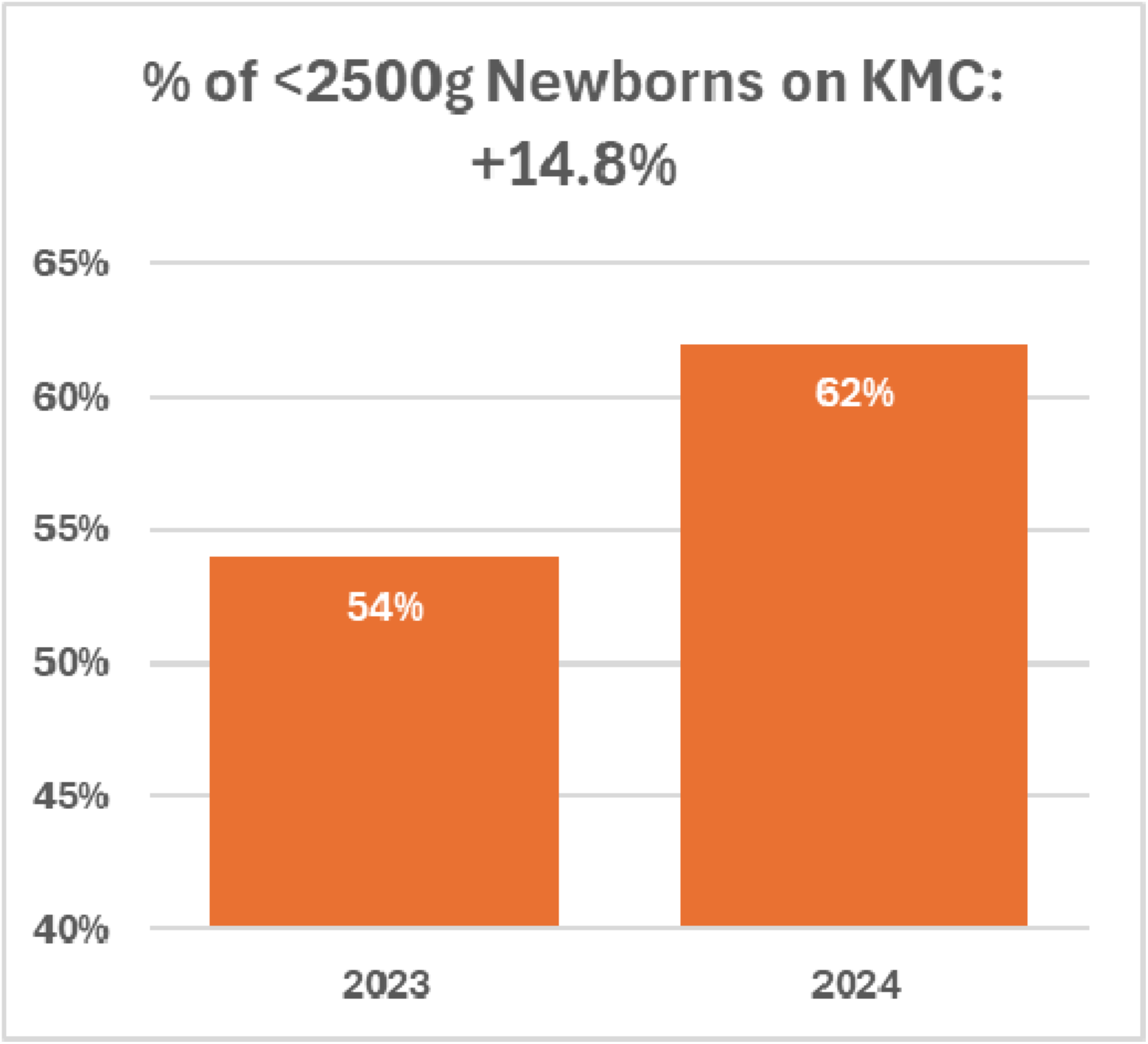
Percentage of newborns weighing less than 2500 grams who were admitted to facilities in Bulamu-supported districts and initiated on Kangaroo Mother Care (KMC) in 2023 and 2024

From an *adoption* standpoint, referrals from Bulamu-supported facilities to those outside the district declined by 7%, suggesting improved local capacity to manage complications. Additionally, 41.3% of supported facilities (45 of 109) reported increased use of KMC for low-birth weight newborns (2500 grams), which is a substantial proportion of supported facilities, but is also an area for continued support.

*Implementation feasibility* was also reflected by a 7% increase in referrals from non-Bulamu facilities to Bulamu-supported facilities, indicating that these sites were increasingly recognized as capable centers for maternal and newborn care.

In terms of *implementation fidelity,* the 2024 coverage of KMC among newborns under 2500 grams reached 62%. We will continue to monitor 2025 data to begin exploring maintenance of these EBIs.

## 4. Discussion

This study evaluated the implementation success of Bulamu’s integrated maternal and newborn health program across eight districts in Uganda. Using the RE-AIM framework, we found that Bulamu’s comprehensive support model-combining workforce training and supply provision was associated with expanded service access, improved clinical capacity, and notable reductions in adverse maternal and newborn outcomes. All supported facilities received the full intervention package, and most showed strong implementation fidelity and maintenance of core program elements. The RE-AIM outcomes suggest that Bulamu’s approach effectively addressed critical health system bottlenecks-such as gaps in clinical training, shortages of essential equipment, and frequent referrals to higher-tier facilities, which contribute to high maternal and neonatal mortality in Uganda and similar settings. Additionally, early evidence for staff training independent of Bulamu programming and increased referrals to Bulamu-supported sites from non-Bulamu-supported sites indicates promise for future sustainability and opportunities for expansion of high-fidelity programming.

Our findings add to the growing body of evidence that integrated, system-level interventions can be both scalable and impact in low-resource contexts. They also highlight the importance of not only implementing evidence-based clinical practices like WHO ENC, and HMS, but pairing them with robust support strategies that enable sustainability and local ownership.

### 4.1 Next steps

To ensure continued progress and sustainability, it is critical to maintain core model activities, including clinical training, transportation support, and the provision of essential medical devices and supplies. A key priority moving forward is to continue equipping district healthcare workers with the knowledge and skills necessary to independently deliver training and mentorship to their peers. This approach, already being implemented in some districts, such as Budaka, has the potential to foster clinician ownership and build long-term sustainability. Strengthening the continuum of maternal and newborn care also remains essential. A comprehensive maternal and newborn health (MNH) program, emphasizing collaboration between obstetric and neonatal services, will be important for advancing high-quality care across the perinatal period.

Ongoing evaluation of the MNH program is vital to refine the model and ensure its effectiveness in improving outcomes. This includes comprehensive reporting of all key EBIs across all participating facilities, enabling appropriate monitoring and timely adjustments to implementation strategies.

### 4.2 Implications

Beyond its programmatic implications, this study advances our understanding of implementing an integrated maternal and newborn program and the pragmatic use of implementation research (IR) tools in resource-limited settings. Our findings contribute to the growing body of literature demonstrating how integrated, facility-based maternal and newborn care interventions can substantially improve health outcomes in low-resource settings. The WHO has advocated for an integrated approach, recognizing that meaningful improvements in newborn health require care that begins during the prenatal period.^14,20^ Similar to prior evaluations of the WHO’s ENC and HMS, we observed a reduction in maternal deaths and increased adoption of evidence-based practices like KMC.^11,14^ However, unlike many single-intervention studies, Bulamu’s model combined District-wide training simultaneous training for all staff, equipment provision, and data-driven quality improvement, aligning with recent calls from global health stakeholders to take a more systems-strengthening approach to maternal and newborn health.^24^ The improvements in both implementation fidelity and EBI coverage suggest that this model may be uniquely positioned to address the persistent barriers— such as the “three delays”—that continue to drive high mortality rates in Uganda and similar settings.^25^

Furthermore, IR tools were effectively leveraged to extract lessons and learnings from routine program implementation. The use of IR allows us to not only evaluate program outcomes but also understand how and why Bulamu’s program performed in the various districts in Uganda. This work contributes to the growing literature surrounding the pragmatic use of IR for monitoring program performance and improving maternal and newborn care.^26,27^ Holtrop et. al. echoes this sentiment through their discussion of the evolution of RE-AIM from a theoretical framework to a practical tool that can support planning and evaluation of health interventions in real time and real-world settings.^22^ This work aligns with current research priorities that emphasize the importance of publishing programmatic findings,^28^ as global health and implementation science are not advancing quickly enough.^29,30^ To support the populations that we serve, we can integrate IR into health programs to generate critical insight and accelerate improvements in care.^30^

## Future research possibilities

Future research should focus on cost-effectiveness analysis, as well as mechanisms for scale-up, and sustainability beyond external support, such as through local ownership. Qualitative studies focused on the perspective of frontline workers and systems-level stakeholders can deepen understanding of implementation facilitators, such as factors driving adoption and long-term impact. Additionally, realist evaluation approaches can help unpack how and why the intervention works in specific contexts. As implementation science evolves, our study reinforces calls for more embedded, systems-informed interventions that are rigorously evaluated across multiple dimensions of impact.^31^

## Limitations

The 2023 data collection at Bulamu-supported sites faced several limitations, particularly in relation to data capture tools, inaccuracies in data entry, and overall data quality. However, it represented the most reliable and traceable data available from the primary data collection tools at the time. In 2024, Bulamu prioritized improving data management by implementing standardized data management tools and practices aimed at addressing these gaps and enhancing data integrity. While the 2023 data had quality concerns, it provided valuable preliminary findings that serve as a foundation for future analyses. Additionally, EBI outcomes that were collected were limited and do not reflect comprehensive outcomes of WHO ENC and HMS. The study’s insights will be further refined and expanded upon with higher-quality data in the coming years. The study was retrospective in nature and not designed as a hybrid trial, which limits its ability to simultaneously assess both the effectiveness of interventions and the processes by which they were implemented. As a result, it does not provide real-time insights into contextual factors or implementation strategies that may have influenced outcomes, reducing the applicability of findings to inform future scale-up or adaptation efforts. Additionally, this study found an increase in institutional perinatal mortality between 2023 and 2024. An increase in the reported number of perinatal deaths is anticipated as service utilization continues to increase, and we strengthen facility-level data collection, and may not reflect true increases in perinatal mortality rate post-Bulamu programming. From a programmatic perspective, our focus will be on using the mortality data to inform the design of strategies aimed at reducing newborn deaths. This may include exploring opportunities such as prioritizing community-based interventions.

## Conclusions

This study demonstrates that an integrated, maternal and newborn health program, centered around WHO recommended trainings and bolstered by pragmatic implementation strategies can drive meaningful improvements in both service delivery and health outcomes in low-resource settings. Through the use of the RE-AIM framework, we documented how Bulamu Healthcare’s model achieved widespread reach, strong adoption, high implementation fidelity, and early signals of sustainability across eight Ugandan Ministry of Health Districts. Notably, the program contributed to reductions in maternal mortality, increased use of KMC, and improved newborn discharge rates. These findings underscore the importance of pairing evidence-based clinical practices with operational supports such as training, supplies, and data-driven

QI. As global health stakeholders call for more integrated and sustainable approaches to maternal and newborn care, this evaluation provides timely, practice-based evidence that such models are both feasible and impactful when implemented with local ownership. Continued investment in systems strengthening and embedded implementation research will be critical to sustaining these gains, informing scale-up, and ultimately reducing preventable maternal and newborn deaths.

## Data Availability

The data used in this study were obtained from country-level District Health Information Software 2 (DHIS2) systems and are not publicly available due to data use agreements and national data protection regulations. Access to these data requires permission from the respective Ministries of Health. Researchers interested in accessing similar data should contact the relevant national health authorities to request permission, subject to applicable policies and procedures. The routinely collected programmatic data is not publicly available but may be available from the corresponding author on reasonable request.

## List of abbreviations

(WHO): World Health Organization
(NMR): Neonatal mortality ratio
(MMR): Maternal mortality ratio
(ENC): Essential newborn care
(HMS): Helping mothers survive
(CPAP): Continuous positive airway pressure
(QI): Quality improvement
(ANC): Antenatal care

## Ethics approval and consent to participate

This study was a program evaluation utilizing de-identified data and did not involve human subjects research as defined by institutional review board (IRB) standards. As such, ethical review and informed consent were not required. All data analyzed were collected as part of routine programmatic activities and were anonymized prior to evaluation to protect participant confidentiality.

## Consent for publication

Not applicable

## Authors’ contributions

This work was done in partnership with Bulamu Healthcare. The author’s views are their own, and not necessarily from any of the institutions they represent. KD, SB, RB, and ZB made substantial contributions to the conception of this work. KD, RB, RS, YV, and AM made significant contributions to the design of this work. KD, AM, RB, YV, and RS made substantial contributions to the acquisition and interpretation of the data. KD drafted the work with substantial revisions from RB, AM, YV, and RS. All authors read and approved the final manuscript and agree to be personally accountable for the author’s own contributions and to ensure that questions related to the accuracy or integrity of any part of the work, even ones in which the author was not personally involved, are appropriately investigated, resolved, and the resolution documented in the literature.

## Acknowledgements

We would like to acknowledge and thank the dedicated health workers who deliver this program, the families that interacted with the Bulamu program, the Ugandan Ministry of Health, and district local governments for their support.

